# The Virtual Cardiologist: Three Deep Learning Pipelines in an Inexpensive Portable Device and Web/Mobile Application for Rapid Cardiovascular Diagnosis and Clinical Decision-Making

**DOI:** 10.1101/2024.05.27.24307981

**Authors:** Shiv Mehrotra-Varma

## Abstract

**Background:** Heart disease is the foremost cause of death worldwide, and specialty care such as cardiologists and expensive Doppler echocardiograms are scarce in rural areas. There is significant variability among physicians in interpreting heart sounds and 2D echocardiograms from more basic, widely available tools like stethoscopes and point-of-care ultrasound devices (POCUS), leading to treatment delays.

**Methods:** 26 Deep-learning models were tested to optimally [a] stratify severity of aortic stenosis from POCUS 2D echocardiogram images [b] estimate cardiac ejection fraction (heart failure) from POCUS 2D echocardiogram videos and [c] identify pathologic heart murmurs using digital stethoscope recordings. The top performing model for each was integrated into a portable, inexpensive Raspberry-Pi device (Virtual Cardiologist), mobile application, and website. Each model was tested on two independent datasets to ensure accuracy. The heart murmur algorithm was also tested on 323 patients by recording their heart sounds via a custom and digital stethoscope in an outpatient setting for clinical validation.

**Results:** The three top-performing models outperformed cardiologists and previous AI methods. The aortic stenosis model (AUC=0.89) utilized EfficientNet, a CNN with compound scaling built on a pre-trained computational backbone. The ejection fraction model (EchoSwin) incorporated Microsoft Swin transformers modified for video input (MAE=5.6). The heart murmur model employed an LSTM Autoencoder algorithm (AUC=0.92).

**Conclusion:** The Virtual Cardiologist can serve as an accurate tool for patient assessment and triage for referral to tertiary care in remote areas with limited specialty care. This is the first tool of its kind for detection of multiple cardiovascular morbidities.

## 1 INTRODUCTION

The CDC states that 46 million Americans and the WHO estimates 3.5 billion people worldwide have limited access to specialty health care services.^1^ There are 46 million ER visits for chest pain annually, and it is the number one cause of death in the US and worldwide, with annual healthcare costs at $370 billion. 6.7% of residents in the Central Valley of California have cardiovascular disease, and 7 million people have limited access to healthcare.

Health care providers commonly have to make urgent clinical decisions with only heart sounds obtained by auscultation with a stethoscope and limited echocardiogram images from relatively inexpensive handheld point-of care ultrasound devices (POCUS). Both modalities require substantial expertise for accurate interpretation.

Multiple studies have shown diagnostic accuracy for auscultation among doctors have eroded significantly (27-68% accuracy).^2^ Point of Care Ultrasound devices can record 2-D Echo images and are often a first line tool to assess cardiac anatomy and function, including wall motion, valve disease and enlargement of heart chambers. A routine echocardiogram takes approximately 10-30 videos and 2000 images, and physicians have extremely limited time to interpret the data. There is substantial variation in human assessment of cardiac function and valve disease with some studies reporting accuracy rates to be 67-88%.^3^

Real-time determination for the need for acute intervention as well as responsible resource allocation can be a considerable challenge, especially in rural emergency settings with limited resources.

The goal of this project was to assist clinicians in remote areas in immediate decision-making where specialty care and highly expensive doppler echocardiogram machines are unavailable, shift the burden of initial diagnosis from advanced echocardiographers and specialists as part of a more cost-effective screening and diagnostic cascade, and identify patients requiring specialty intervention at earlier stages of disease to facilitate early intervention, thereby reducing mortality rates and decreasing healthcare costs by reducing the need for ICU stays.

We designed and tested *numerous* (26) AI models to determine the most accurate method to identify the presence of aortic stenosis (AS), the most common valvular heart disease, from limited 2D echocardiogram images, determine the cardiac ejection fraction and the presence of heart failure from an Echocardiogram video, and identify and classify heart murmurs from recorded heart sounds.

All models underwent a two-step verification process to ensure accuracy. 2-D Echocardiogram images and videos are readily available from inexpensive Point of Care Ultrasound Devices (POCUS). A digital stethoscope was designed to capture heart murmur recordings.

Finally, a portable handheld device containing a Raspberry-Pi (The Virtual Cardiologist), a mobile application, and a PC application were designed to store each of the three top performing models, capable of receiving and analyzing an input from either a POCUS or digital stethoscope.

## 2 RESEARCH DESIGN AND METHODS

### 2.1 Automated Aortic Stenosis Classification (15 Models)

The first goal was to develop and test an accurate model to identify and stratify the severity of aortic stenosis from limited 2-D echo views. The de-identified Tufts Medical Center TMED-2^4^ dataset (599 patient studies verified by expert cardiologists) was used for training (80% of data) and testing (20% of data). Our methodology required a GPU to explore a comprehensive array of hyperparameters, laying the groundwork for testing across a spectrum of ML models. To identify the model with highest accuracy, the evaluation included several types of convolutional neural networks (CNNs). Furthermore, the effects of fine-tuning strategies were evaluated to refine and adapt pre-trained models to align more closely with the requirements presented by our data and address unique challenges in the context of medical imaging analysis. The best model was reported after training from scratch and fine-tuning on ImageNet-21k weights to find the model with highest accuracy.

Accurate classification of aortic stenosis requires integration of structural and hemodynamic parameters in multiple Echo images. 15 deep learning models were trained and tested to determine the most accurate computational architecture for the detection and classification of AS (**Figure 1**). This was done by the following 3 steps: [a] Echo view Identification [b] Identification of presence and severity of AS (per American Society of Echocardiography guidelines^5^) [c] Prioritized weighting of PLAX (parasternal long axis) and PSAX (parasternal short axis) Aortic valve views as stronger evidence of disease severity to make a final prediction.

**Figure 1.**
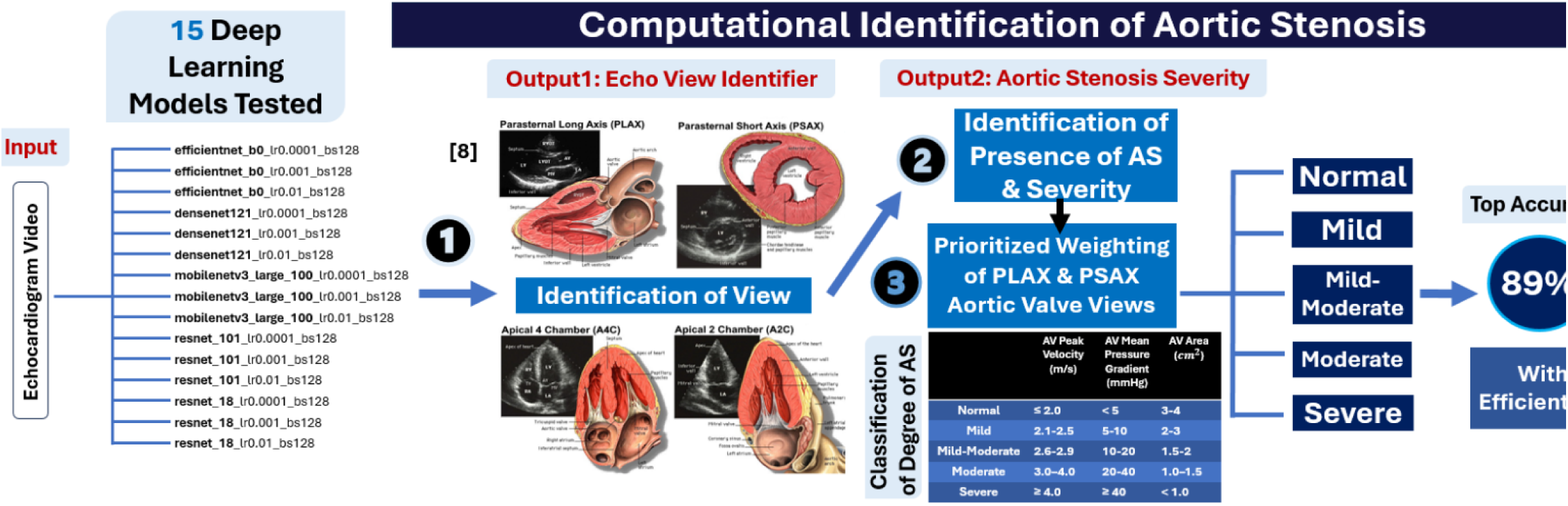
Computational Identification of Aortic Stenosis. Fifteen different deep learning models were trained to identify the type of view and presence and severity of aortic stenosis from an inputted echocardiogram video. American Society of Echocardiography guidelines were used as classification of degree of aortic stenosis from AV (Aortic Valve) Peak Velocity, AV Mean Pressure Gradient, and Aortic Valve Area. The highest accuracy was achieved with an EfficientNet model.

### 2.2 Automated Estimation of Cardiac Ejection Fraction (6 Models)

The second goal was to develop a Deep Learning model for the intricate task of identifying cardiac ejection fraction from video data. This required integration of a recurrent component into an existing Microsoft Swin Transformers model to accommodate the temporal characteristics inherent in video data, which differ significantly from static images. The experimental framework encompassed testing a diverse range of machine learning models on a GPU, including Recurrent Neural Networks and Transformer-based architectures, which are particularly adept at handling sequential data. The performance of these models was meticulously evaluated, considering both training from scratch and fine-tuning using pre-existing weights, such as those from ImageNet-21k.

To determine Ejection Fraction accurately, 6 deep learning models were tested (Figure 2). The training (80% of the data) and testing (20% of the data) was done using the publicly available dataset from Stanford Medical Center called EchoNet-Dynamic, which was labeled and cross checked by expert cardiologists.^6^

**Figure 2.**
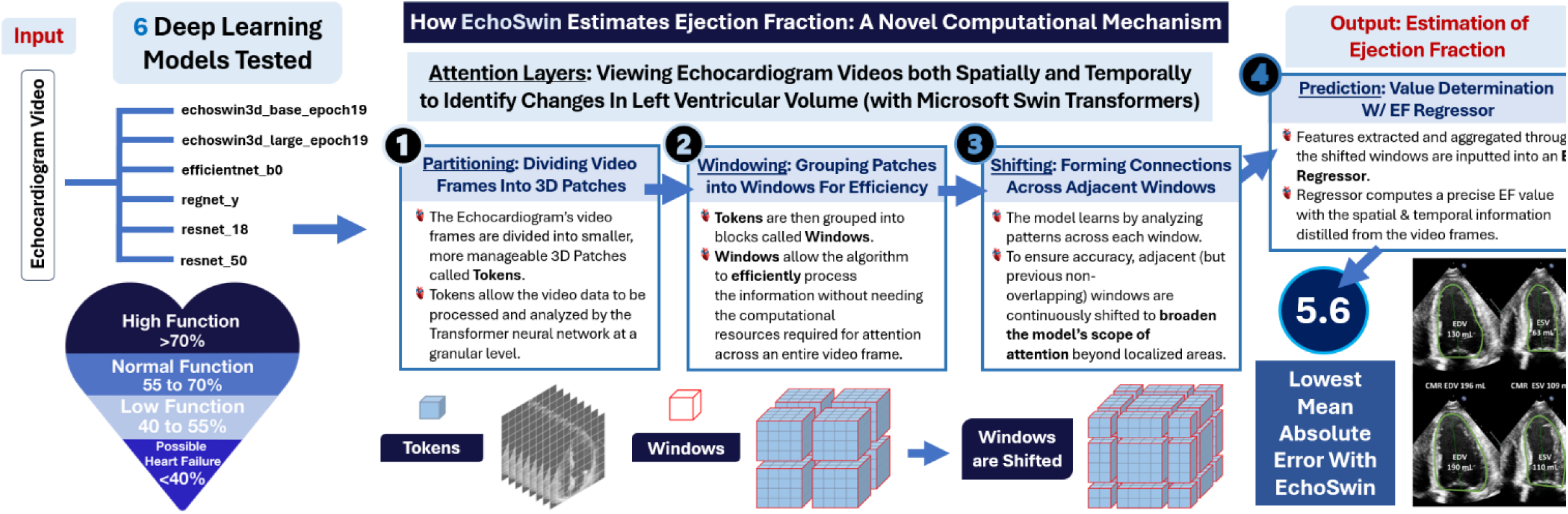
Estimation of Ejection Fraction. Six different deep learning models were trained to estimate ejection fraction from an echocardiogram video input to identify likelihood of heart failure. The highest accuracy was achieved with a modified EchoSwin model, which identifies changes in left ventricular volume over three steps: partitioning, windowing, and shifting.

The primary model utilized Microsoft Hierarchical Swin Transformers^7^ (used for static image analysis) that was modified to receive an input video. The model has a features extractor that determines End Systolic and End Diastolic Volume from one complete cardiac cycle (heartbeat) and creates a features map. It then views the data both spatially and temporally, and finally an EF Regressor processes the information to output a scalar value for EF.

### 2.3 Automated Heart Murmur Detection (5 Models)

The third goal was to develop an inexpensive electronic stethoscope that converts heart sounds to analog voltage, which is amplified and filtered to reduce ambient noise and digitally processed for murmur identification. The experimental design was to train and test 5 different AI models on data compiled from publicly available heart sound recordings annotated by expert cardiologists and by echocardiograms from University of Michigan School of Medicine^8^, University of Washington School of Medicine^9^, etc (Figure 3).

**Figure 3.**
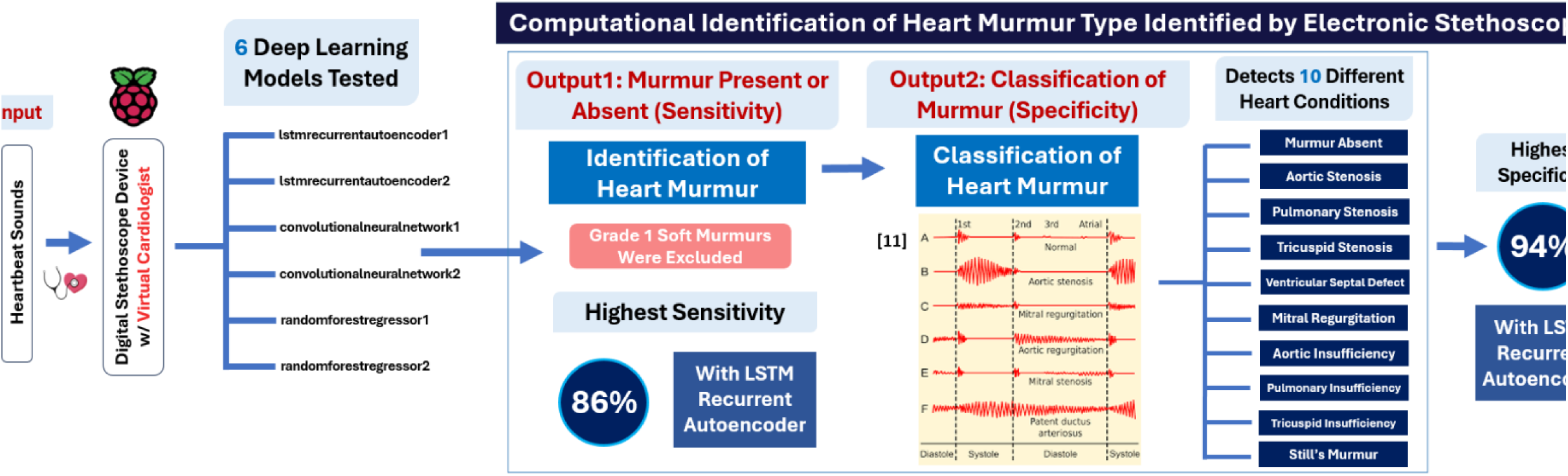
Identification of Heart Murmur Type. Six different deep learning models were trained to detect and classify heart murmurs from inputted heartbeat sounds, recorded with a custom electronic stethoscope that can connect to the Virtual Cardiologist device. The highest accuracy was achieved with an LSTM Recurrent Autoencoder model.

Developing a way to accurately identify heart murmurs required training (using 80% of data) and testing (using 20% of data) of 5 different Deep Learning models on approximately 1000 annotated heart recordings to identify the most accurate model. An inexpensive electronic stethoscope was made to record heart sounds, which were processed through the most accurate Deep Learning (DL) algorithm (LSTM Recurrent Autoencoder) to output the murmur type and results on the Virtual Cardiologist device.

The models were trained to detect ten different conditions: Murmur Absent, Aortic Stenosis, Pulmonary Stenosis, Tricuspid Stenosis, Ventricular Septal Defect, Mitral Regurgitation, Pulmonary Insufficiency, Tricuspid Insufficiency, and Still’s Murmur. Accuracy of the model was tested on the dataset and heart sound recordings from 323 patients at a Fresno outpatient clinic.

### 2.4 Two-Fold Verification Testing

Accuracy for each model was measured with two-fold verification testing (80/20 Train Test Split), including in a clinical setting (Primary Care clinic) for heart murmurs (Figure 4). For ease of comparison, results were then averaged across the two datasets. For the second dataset for aortic stenosis, the EchoNet dataset did not contain labels for AS, so the database could only be used to measure the accuracy of the Echo ViewFinder. Moreover, besides EchoNet, no public dataset labeled for ejection fraction videos exists. Echo recordings were compiled from various sources to make Dataset 2 for EF. Finally, no publicly available dataset labeled for all ten conditions of heart murmurs exists, so Dataset 1 for murmurs was compiled from a variety of sources (U. Michigan, U Washington School of Medicine). To test the accuracy of the model, Dataset 2 for murmurs was recorded and tested using a custom digital stethoscope to obtain heart sounds of 323 anonymous patients from a local outpatient clinic in Fresno.

**Figure 4.**
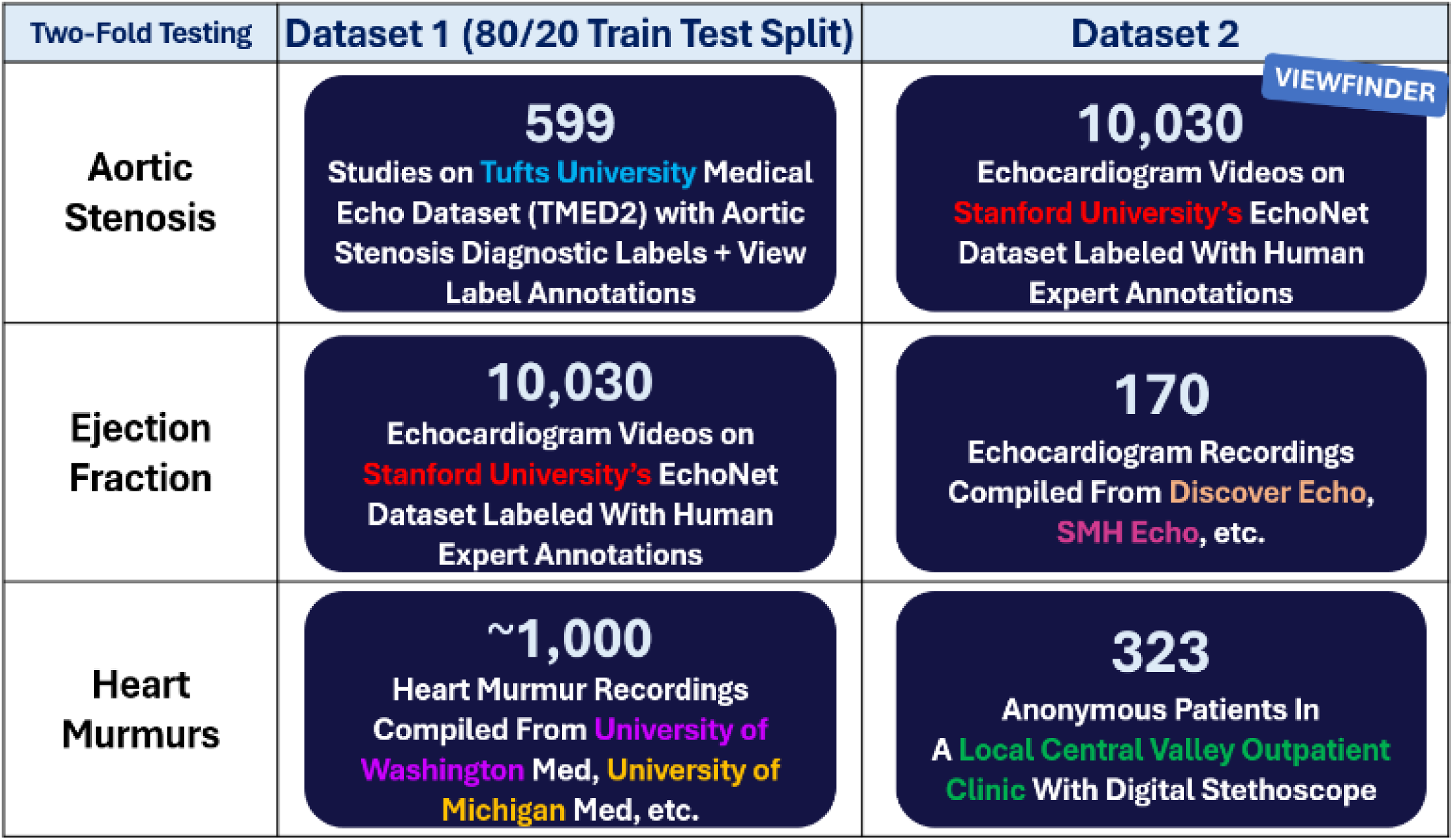
All 26 models were tested on two independent datasets.

### 2.5 Model Validation in A Local Outpatient Clinic

I obtained informed consent from the patient or guardian, then recorded the patient’s heart sounds using an electronic stethoscope. Recorded heart sounds were de-identified and assigned a numeric id. They were processed by the top performing model on a raspberry pi device for murmur classification. Clinic medical staff annotated the recordings using the assigned numeric ID with previously determined diagnoses, thereby maintaining anonymity of the recording. Ground truth diagnosis was based on results of a doppler echocardiogram report by an experienced cardiologist. The model results were cross checked with the annotation from the patient’s medical record for accuracy.

### 2.6 Validation Metrics

AUC and Loss were extracted for each of the sixteen aortic stenosis models. For the most accurate model, sensitivity (ability to designate an individual with a disease as positive), specificity (ability to designate an individual who does not have a disease as negative), and ViewFinder accuracy were also extracted.

For EF, it was not possible to extract area under the curve for the EchoSwin models due to the unique nature of the Swin architecture. Instead, Loss, Mean Absolute Error (MAE), Coefficient of Determination (R^2), and Root Mean Squared Error (RMSE) were compared across models as measures of performance.

For Heart Murmurs, AUC and Loss were extracted for each of the six models (averaged across all ten conditions). For the most accurate model, specificity and sensitivity were also extracted. For testing at the local outpatient clinic, the number of patients correctly classified was divided by the actual number of patients diagnosed for each of the ten conditions.

## 4 RESULTS

Efficient Net, with a learning rate of 0.001 and a batch size of 128, was the most accurate of the sixteen models, with an AUC of 0.89, sensitivity of 0.90, and specificity of 0.89. (Table 1, Figure 5).

**Tables 1-3.**
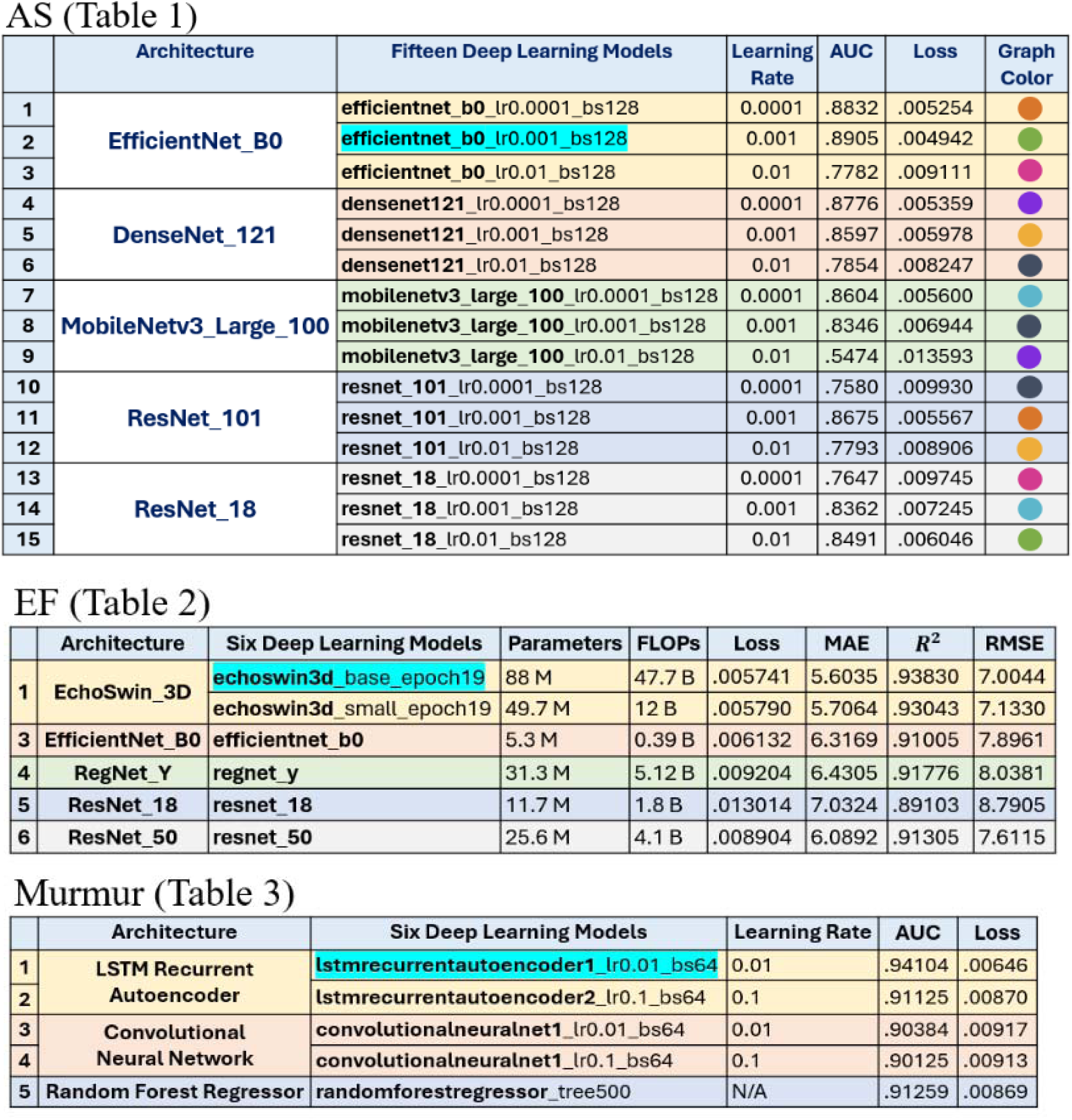
Results for 15 models to detect and classify AS (left), 6 models to estimate EF (top right), and 5 models to detect and classify heart murmurs (bottom right). The following were obtained and reported-AUC and Loss values for the AS models, Loss, MAE, MSE, R2, and RMSE values for the EF models, and AUC and Loss values for the Murmur models. Table 1 for AS contains the legend for Figure 5.

|  | Architecture | Fifteen Deep Learning Models | Learning Rate | AUC | Loss | Graph Color |
| --- | --- | --- | --- | --- | --- | --- |
| 1 | EfficientNet_B0 | efficientnet_b0_lr0.0001_bs128 | 0.0001 | .8832 | .005254 |  |
| 2 |  | efficientnet_b0_lr0.001_bs128 | 0.001 | .8905 | .004942 |  |
| 3 |  | efficientnet_b0_lr0.01_bs128 | 0.01 | .7782 | .009111 |  |
| 4 | DenseNet_121 | densenet121_lr0.0001_bs128 | 0.0001 | .8776 | .005359 |  |
| 5 |  | densenet121_lr0.001_bs128 | 0.001 | .8597 | .005978 |  |
| 6 |  | densenet121_lr0.01_bs128 | 0.01 | .7854 | .008247 |  |
| 7 | MobileNetv3_Large_100 | mobilenetv3_large_100_lr0.0001_bs128 | 0.0001 | .8604 | .005600 |  |
| 8 |  | mobilenetv3_large_100_lr0.001_bs128 | 0.001 | .8346 | .006944 |  |
| 9 |  | mobilenetv3_large_100_lr0.01_bs128 | 0.01 | .5474 | .013593 |  |
| 10 | ResNet_101 | resnet_101_lr0.0001_bs128 | 0.0001 | .7580 | .009930 |  |
| 11 |  | resnet_101_lr0.001_bs128 | 0.001 | .8675 | .005567 |  |
| 12 |  | resnet_101_lr0.01_bs128 | 0.01 | .7793 | .008906 |  |
| 13 | ResNet_18 | resnet_18_lr0.0001_bs128 | 0.0001 | .7647 | .009745 |  |
| 14 |  | resnet_18_lr0.001_bs128 | 0.001 | .8362 | .007245 |  |
| 15 |  | resnet_18_lr0.01_bs128 | 0.01 | .8491 | .006046 |  |

EF (Table 2)
|  | Architecture | Six Deep Learning Models | Parameters | FLOPs | Loss | MAE | R<sup>2</sup> | RMSE |
| --- | --- | --- | --- | --- | --- | --- | --- | --- |
| 1 | EchoSwin_3D | echoswin3d_base_epoch19 | 88 M | 47.7 B | .005741 | 5.6035 | .93830 | 7.0044 |
|  |  | echoswin3d_small_epoch19 | 49.7 M | 12 B | .005790 | 5.7064 | .93043 | 7.1330 |
| 3 | EfficientNet_B0 | efficientnet_b0 | 5.3 M | 0.39 B | .006132 | 6.3169 | .91005 | 7.8961 |
| 4 | RegNet_Y | regnet_y | 31.3 M | 5.12 B | .009204 | 6.4305 | .91776 | 8.0381 |
| 5 | ResNet_18 | resnet_18 | 11.7 M | 1.8 B | .013014 | 7.0324 | .89103 | 8.7905 |
| 6 | ResNet_50 | resnet_50 | 25.6 M | 4.1 B | .008904 | 6.0892 | .91305 | 7.6115 |

Murmur (Table 3)
|  | Architecture | Six Deep Learning Models | Learning Rate | AUC | Loss |
| --- | --- | --- | --- | --- | --- |
| 1 | LSTM Recurrent Autoencoder | lstmrecurrentautoencoder1_lr0.01_bs64 | 0.01 | .94104 | .00646 |
| 2 |  | lstmrecurrentautoencoder2_lr0.1_bs64 | 0.1 | .91125 | .00870 |
| 3 | Convolutional Neural Network | convolutionalneuralnet1_lr0.01_bs64 | 0.01 | .90384 | .00917 |
| 4 |  | convolutionalneuralnet1_lr0.1_bs64 | 0.1 | .90125 | .00913 |
| 5 | Random Forest Regressor | randomforestregressor_tree500 | N/A | .91259 | .00869 |

**Figure 5.**
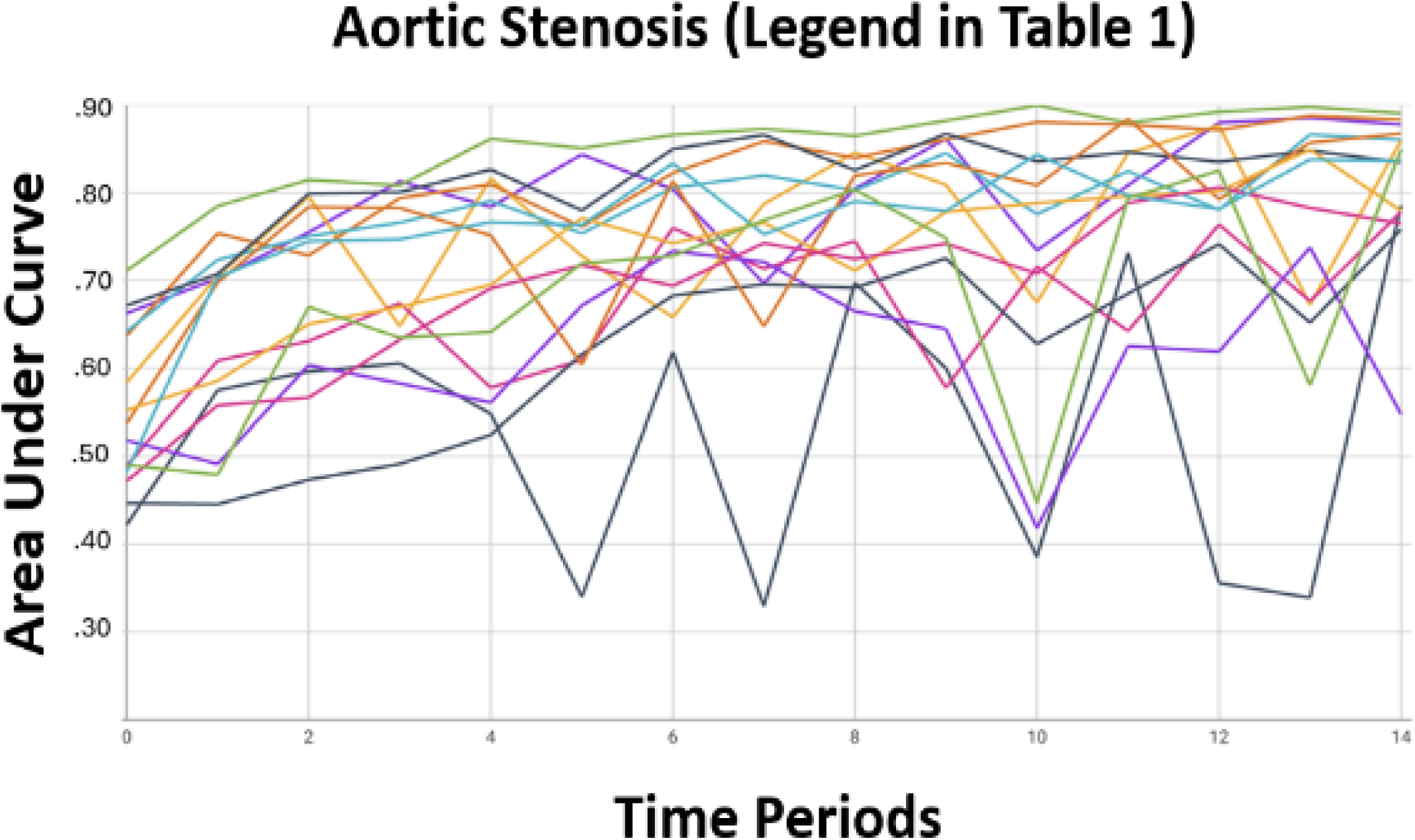
AS AUC

EchoSwin base, with 49.7 M parameters (in log scale), was the most accurate model for EF (Table 2).

LSTM Recurrent Autoencoder, with a learning rate of 0.01, was the most accurate of the six models tested, with an AUC of 0.92, sensitivity of 0.94, and specificity of 0.86 (Table 3, Figure 6). Table 4 shows the results of clinical validation testing.

**Figure 6.**
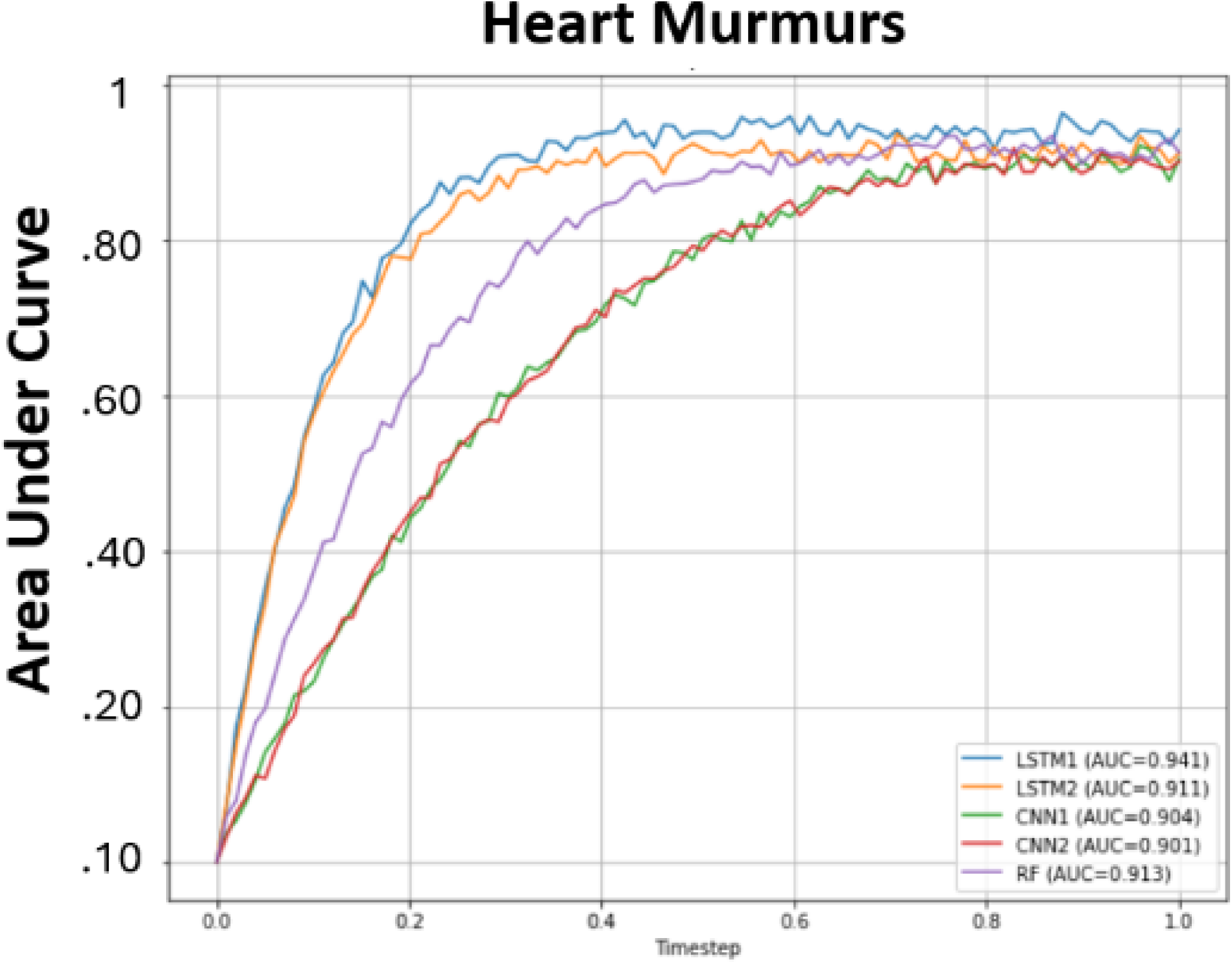
Murmurs AUC

**Table 4.** 323 patients were tested at a local outpatient clinic. Model results were compared with ground truth diagnosis.

|  | **Diagnosis** | **# Pts With Condition** | **# Pts Correctly Classified** |
| --- | --- | --- | --- |
| **1** | **Murmur Absent** | **266** | **266 (100%)** |
| **2** | **Aortic Stenosis** | **4** | **3 (75%)** |
| **3** | **Pulmonary Stenosis** | **1** | **1 (100%)** |
| **4** | **Tricuspid Stenosis** | **0** | **-** |
| **5** | **Ventricular Septal Defect** | **33** | **28 (84.85%)** |
| **6** | **Mitral Regurgitation** | **3** | **3 (100%)** |
| **7** | **Aortic Insufficiency** | **1** | **1 (100%)** |
| **8** | **Pulmonary Insufficiency** | **0** | **-** |
| **9** | **Tricuspid Insufficiency** | **2** | **2 (100%)** |
| **10** | **Still's Murmur** | **14** | **13 (92.86%)** |

## 5 DISCUSSION

The greatest benefit of this prototype is the ability for healthcare providers with limited access to specialty services to have a single, inexpensive, accurate, and portable device with the capacity to identify several common cardiac pathologies to assist in rapid clinical decision-making. Incorporation of the ability to process both 2-D echo images and videos, as well as heart sound recordings from auscultation via a stethoscope creates a foundation for a platform that may allow comprehensive cardiac evaluation of a patient with accuracy metrics approaching that of an experienced cardiologist. Additional research to expand the diagnostic capability of the device and clinical testing on a diverse patient population is essential to ensure reliability.

### 5.1 Aortic Stenosis

The algorithm to detect Aortic stenosis is particularly effective in preventing missed cases (very high negative predictive value) and has the unique capability of stratifying severity. It is an ideal tool for cost-effective screening in patients exhibiting mild or no symptoms, thereby enabling early diagnosis, improved outcomes, and reduction in healthcare costs.

Limitations of the model include underrepresentation of nonwhite patients in the training cohort, although the echocardiography-based imaging diagnosis of AS should not have any biological differences on the basis of race. Other limitations are a relatively modest positive predictive value, and the absence of methods to integrate AI algorithm results into standard electronic medical records.

The EfficientNet model likely demonstrated higher accuracy compared to other model types for two reasons: the use of a pretrained model backbone *and* the use of a novel compound scaling technique.

Unlike a traditional CNN, the model was first pretrained on the extensive ImageNet dataset to develop a strong visual backbone for identifying features in images, in a process called transfer learning. This pretrained model was then fine-tuned on echo videos, making it more receptive to the nuances of echocardiogram images. The use of transfer learning allowed the model to learn robust generalizable features before specializing to the target task of AS identification.

EfficientNet also utilized compound scaling to produce a stronger model backbone. Previous research has demonstrated that bigger DL networks with larger width, depth, or resolution tend to achieve higher accuracy, but the accuracy gain quickly saturates after reaching 80%. This is limiting as traditional neural network scaling practices adjust width, depth, and resolution in isolation. For example, when only scaling network width without changing depth and resolution, the backbone’s accuracy saturates quickly. Compound scaling was applied to the EfficientNet model to scale dimensions jointly rather than independently. With deeper and higher resolution, compound scaling achieves much better accuracy with the same computational cost compared to a traditional neural network that uses independent scaling.

### 5.2 Cardiac Ejection Fraction

In terms of Ejection Fraction (EF) measurement, the EchoSwin achieved a 5.6 Mean Absolute Error, matching or exceeding the performance of human echocardiographers and existing AI methods. Further improvement of Echo-swin performance can be attempted by aggregating the features extraction at every stage before being processed by the EF regressor. Also, ground truth values for testing for model accuracy in the datasets used were labeled by expert cardiologists; however inter-observer inaccuracies are well described. Cardiac MRI is considered the most accurate way to assess EF, and if databases labeled with values cross-referenced by cardiac MRI become available in the future, better accuracy metrics can be obtained. EchoSwin has a unique method of analyzing echocardiogram videos spatio-temporally in four steps rather than just training the model to recognize patterns like in a traditional CNN, which likely led to its higher accuracy. Microsoft Swin Transformer models were modified to analyze **c**ardiac ultrasound video containing left ventricular motion in the following steps: [1] echo frames were pre-processed into a fixed length sequence to be used as training data. [2] a hierarchy of Swin transformers were implemented to a single Transformers Encoders (TE) module to extract spatio-temporal features from the input video frames. [3] The transformers use the shifted window attention technique (Figure 2) to capture both local and global context. [4] A regressor module processes the Swin features maps from the TE module and generates a scalar EF value as output. It applies several linear layers to condense the features into a single EF prediction.

### 5.3 Heart Murmurs

Murmur detection by the device outperformed both internal medicine doctors and experienced cardiologists and was validated in a clinical setting with 323 patients.

The most accurate model was an LSTM Recurrent Autoencoder, which was modified to analyze heart sound recordings over two LSTM layers. The first LSTM layer with 128 hidden units condenses the 1500 timestep input down to a 128-dimension representation. This is further compressed to 64 dimensions by the second LSTM layer. Decoder LSTM layers then expand this 64-dimension encoding back to the original sequence length in order to reconstruct the input. A final linear layer attempts to accurately rebuild the input heartbeat pattern. The model was trained over 140 epochs and optimized through an Adam optimizer and L1 loss function. It flags potentially anomalous inputs by checking if the reconstruction error exceeds a threshold level (novel threshold equation: **2**(-mismatch[0]/thresholds[name])** determined during training.

Auscultation skills of physicians have declined substantially, especially in the era of electronic medical records and increasing reliance on technology, making reliable AI augmented auscultation an urgent imperative for patient care. Challenges include the model’s inability to recognize lung sounds such as wheezes and crackles which are easily identified by physicians and can add artifact error to the signal. This can impact the accuracy/ classification ability of the algorithm. A possible field for further research can be to make a Machine Learning tool to accurately identify both heart and lung sounds. The algorithm was tested on patients in an outpatient primary care office where the number of patients with certain types of murmurs/diagnoses was limited. Further testing needs to be done on larger groups of patients to assess accuracy of murmur identification in diverse ethnicities, ages and pathologies.

Several attempts were made to obtain access to Cardiac Auscultatory Recording Database (CARD) from Johns Hopkins Medical Center for additional testing for murmur classification.^9^ However, at this time, the request is still undergoing review by their utilization committee. If access to CARD is granted, further accuracy testing can be performed in the future.

### 5.4 Further Research

Future research directions include creating deep learning (DL) models to identify a comprehensive range of heart conditions via point-of-care echocardiograms, such as pericardial effusion and other valvular heart diseases. The greatest limiting factor to accomplish this is the availability of accurately labeled datasets which are required for training and testing of AI algorithms. Large academic institutions should be encouraged to allow public access to labeled data scrubbed for patient identification for evaluation of different ML models. This may allow innovation of new architectures better suited for the specific requirements of each diagnostic challenge. Clinical validation in multiple settings that include patients of diverse ages, ethnicities and clinical presentations is the cornerstone of any testing for proof of validity and is essential in the future.

## 6 CONCLUSIONS

An inexpensive portable device using Machine Learning algorithms can provide an accurate tool for patient assessment and triage for referral to tertiary care in settings where specialty services are limited. Research in this field is ongoing, and performance metrics may continue to improve as more labeled data becomes available. Additional research is essential to create AI algorithms that expand diagnostic capabilities to allow for comprehensive cardiovascular evaluation. This prototype has the potential to decrease the profound disparities in treatment of patients in underserved rural areas and ethnic minorities. Real-world usability and clinical validation are crucial in evaluating the effectiveness of these AI applications in healthcare and are a target for future research.

## Data Availability

All data produced in the present study are available upon reasonable request to the authors.

https://tmed.cs.tufts.edu/tmed_v2.html

https://echonet.github.io/lvh/#:~:text=The%20EchoNet%2DLVH%20dataset%20contains,clinical%20care%20at%20Stanford%20Medicine.

https://physionet.org/content/circor-heart-sound/1.0.3/

https://www.med.umich.edu/lrc/psb_open/html/repo/primer_heartsound/primer_heartsound.html

https://depts.washington.edu/physdx/heart/demo.html

## Notes

### Competing Interest Statement

The authors have declared no competing interest.

### Funding Statement

This study did not receive any funding.

### Author Declarations

Ethics committee/IRB of California State University, Fresno gave ethical approval for this work.

